# The Role of Cytokines in Acute and Chronic Postsurgical Pain in Pediatric Patients after Major Musculoskeletal Surgeries

**DOI:** 10.1101/2024.03.27.24304974

**Authors:** Vidya Chidambaran, Qing Duan, Valentina Pilipenko, Susan M. Glynn, Alyssa Sproles, Lisa J. Martin, Michael J. Lacagnina, Christopher D. King, Lili Ding

**Author notes:** **Corresponding author:** Vidya Chidambaran, MD, MS, Professor, Department of Anesthesia, Cincinnati Children’s Hospital Medical Center 3333 Burnet Avenue, Cincinnati, OH, 45229, USA,**, **. **Disclosures:** Research reported in this manuscript was supported by National Institute of Arthritis and Musculoskeletal and Skin Diseases under award number 5R01AR075857 (PI:Chidambaran). The content is solely the responsibility of the authors and does not necessarily represent the official views of the National Institutes of Health.

## Abstract

**Study Objective:** To determine if baseline cytokines and their changes over postoperative days 0-2 (POD0-2) predict acute and chronic postsurgical pain (CPSP) after major surgery.

**Design:** Prospective, observational, longitudinal nested study.

**Setting:** University-affiliated quaternary children’s hospital.

**Patients:** Subjects (≥8 years old) with idiopathic scoliosis undergoing spine fusion or pectus excavatum undergoing Nuss procedure.

**Measurements:** Demographics, surgical, psychosocial measures, pain scores, and opioid use over POD0-2 were collected. Cytokine concentrations were analyzed in serial blood samples collected before and after (up to two weeks) surgery, using Luminex bead arrays. After data preparation, relationships between pre- and post-surgical cytokine concentrations with acute (% time in moderate-severe pain over POD0-2) and chronic (pain score>3/10 beyond 3 months post-surgery) pain were analyzed. After adjusting for covariates, univariate/multivariate regression analyses were conducted to associate baseline cytokine concentrations with postoperative pain, and mixed effects models were used to associate longitudinal cytokine concentrations with pain outcomes.

**Main Results:** Analyses included 3,164 measures of 16 cytokines from 112 subjects (median age 15.3, IQR 13.5-17.0, 54.5% female, 59.8% pectus). Acute postsurgical pain was associated with higher baseline concentrations of GM-CSF (β=0.95, SE 0.31; *p*=.003), IL-1β (β=0.84, SE 0.36; *p*=.02), IL-2 (β=0.78, SE 0.34; *p*=.03), and IL-12 p70 (β=0.88, SE 0.40; *p*=.03) and longitudinal postoperative elevations in GM-CSF (β=1.38, SE 0.57; *p*=.03), IFNγ (β=1.36, SE 0.6; *p*=.03), IL-1β (β=1.25, SE 0.59; *p*=.03), IL-7 (β=1.65, SE 0.7, *p*=.02), and IL-12 p70 (β=1.17, SE 0.58; *p*=.04). In contrast, CPSP was associated with lower baseline concentration of IL-8 (β= -0.39, SE 0.17; *p*=.02), and the risk of developing CPSP was elevated in patients with lower longitudinal postoperative concentrations of IL-6 (β= -0.57, SE 0.26; *p*=.03), IL-8 (β= - 0.68, SE 0.24; *p*=.006), and IL-13 (β= -0.48, SE 0.22; *p*=.03). Furthermore, higher odds for CPSP were found for females (*vs.* males) for IL-2, IL-4, IL-5, IL-6, IL-8, IL-10, and TNFα, and for pectus (*vs.* spine) surgery for IL-8 and IL-10.

**Conclusion:** We identified pro-inflammatory cytokines associated with increased acute postoperative pain and anti-inflammatory cytokines associated with lower CPSP risk, with potential to serve as predictive and prognostic biomarkers.

## 1. Introduction

Major musculoskeletal surgeries are often accompanied by acute postsurgical pain, which can have deleterious effects on long-term function and recovery, leading to chronic postsurgical pain (CPSP). Pectus excavatum repair and spine deformity correction surgery are two such painful surgeries adolescents undergo and are associated with extensive dissection, muscle derangements, inflammation, nerve sensitization [1] and CPSP [2, 3]. Surgeries are often associated with a host of immune responses following tissue injury and incision [4]. These include the release of cytokines, stress responses and acute phase proteins, which can be harmful when excessive. [5] At the same time, these responses are the body’s defense against infection and tissue damage, and their release can be protective. Thus, a simple anti-inflammatory strategy for example with Non-steroidal Anti-inflammatory Drugs raises concerns regarding adverse effects, such as infection and delayed wound-healing[6]. Understanding how these inflammatory responses impact pain outcomes is therefore important to know which cytokines may promote healing and which ones are associated with harmful pain.

Cytokines, as pivotal regulators of immune and inflammatory processes, have emerged as key players in the modulation of pain pathways.[7] Cytokines are soluble molecular substances produced by inflammatory cells. Broadly, they can be either pro-inflammatory (e.g., interleukins (IL)-1, 2, 8, 12; tumor necrosis factor α (TNFα), interferon γ (IFNγ)) or anti-inflammatory (e.g. IL-4, IL-10, IL-13, transforming growth factor beta (TGFβ)), while some such as IL-6 can be both pro- and anti-inflammatory. Pro-inflammatory cytokines may be associated with postoperative pain after musculoskeletal surgeries [8] and may activate neural circuits to maintain hyperalgesia states [9]. Certain inflammatory profiles in the serum and body fluids have been suggested to be characteristic of poorly resolving pain after joint surgery [10]. However, other studies have refuted these findings and did not find associations between perioperative cytokine concentrations with postoperative pain following total knee arthroplasty [11]. Thus, there is a lack of understanding of the role of cytokines in acute and chronic postoperative pain. This is crucial for developing targeted and effective pain management strategies, as a precise understanding of the cytokine response to surgical trauma may inform interventions that would optimize perioperative care, decrease morbidity and enhance recovery.

In this study, we aimed to investigate whether baseline cytokine profiles and longitudinal concentration changes in cytokines after surgery predicted acute and chronic postoperative pain outcomes. We hypothesized pro-inflammatory cytokines such as interferon gamma interferon (IFNγ), IL1β, IL2, IL8, and TNF-α will be associated with increased risk, while anti-inflammatory cytokines such as IL4, IL10, and IL13 will decrease risk for poor pain outcomes after surgery.

## 2. Materials and Methods

This prospective, observational, longitudinal study was nested within a larger genomics study (<u>ClinicalTrials.gov</u> Identifier: NCT02998138, NCT01839461, NCT01731873) and was conducted at a university-affiliated quaternary children’s hospital. The Cincinnati Children’s Institutional Review Board approved the study. Results pertaining to psychosocial factors and epigenetic factors influencing CPSP have been previously published [3, 12, 13]. Written informed consent was obtained from parents and assent from children before enrollment, either on paper or electronically, through Research Electronic Data Capture (REDCap), a secure web application for building and managing online surveys and databases. This study adhered to the Reporting of Observational Studies in Epidemiology (STROBE) guidelines [14].

### 2.1 Participants

Healthy children aged 8 years and above, regardless of sex or race, were included if they had a diagnosis of idiopathic scoliosis/kyphosis or pectus excavatum, American Society of Anesthesiologists physical status ≤ 2 (mild systemic disease) and were scheduled for elective spine fusion or Nuss procedure. They were screened from operating room schedules and recruited if they satisfied inclusion/exclusion criteria. Exclusion criteria included a history of opioid use in the past six months, liver and renal disease, pregnant or breastfeeding females, developmental delay, cancer, and those not fluent in written and/or spoken English. Perioperative management included standardized surgery-specific anesthesia and multimodal pain protocols.

Patients undergoing either surgery received acetaminophen and ketorolac (on postoperative day (POD)0-2), dexamethasone (intraoperative only), muscle relaxants and opioids. Pectus patients also received regional anesthesia, methadone and pregabalin.

### 2.2 Data collection

Data were collected by chart review and via questionnaires administered through REDCap via email. Before surgery, the following data were collected: demographics (sex, age, race), weight, pain scores (numerical rating scale (NRS) 0-10)[15], diagnosis and surgery type. The Pediatric Pain Screening Tool (PPST) was administered preoperatively – a brief, 9-item self-report questionnaire developed for rapid identification of risk for poor pain coping. It has been validated in patients with chronic pain and within this surgical cohort for CPSP risk stratification [12, 16]. The first four items evaluate physical symptoms, and the next four questions evaluate psychosocial constructs. PPST total scores range from 0 to 9. Perioperative data collected included duration of surgery, pain scores (recorded every four hours) and analgesia (opioid medications/doses, (intravenous, oral and patient-controlled analgesia) over 48 hours after surgery. After hospital discharge, questionnaires were administered via REDCap 3-6 and 10–12 months after surgery to obtain pain measures (average and maximum pain scores over the last week).

#### 2.2.1 Blood draws

Blood was collected from intravenous catheters placed for clinical purposes and /or indwelling lines. An 8 ml sample was collected in purple top/EDTA tubes at baseline. Additional samples (1-2 ml) were collected at the scheduled time-points (**Figure 1**). The total amount of blood drawn per patient was approximately 30 ml. Whole blood samples were centrifuged at 3500 rpm for 10 minutes, plasma aliquoted into two tubes (minimal volume per freezer aliquot tube: 250 microliters) and stored at -80°C until analysis.

**Figure 1:**
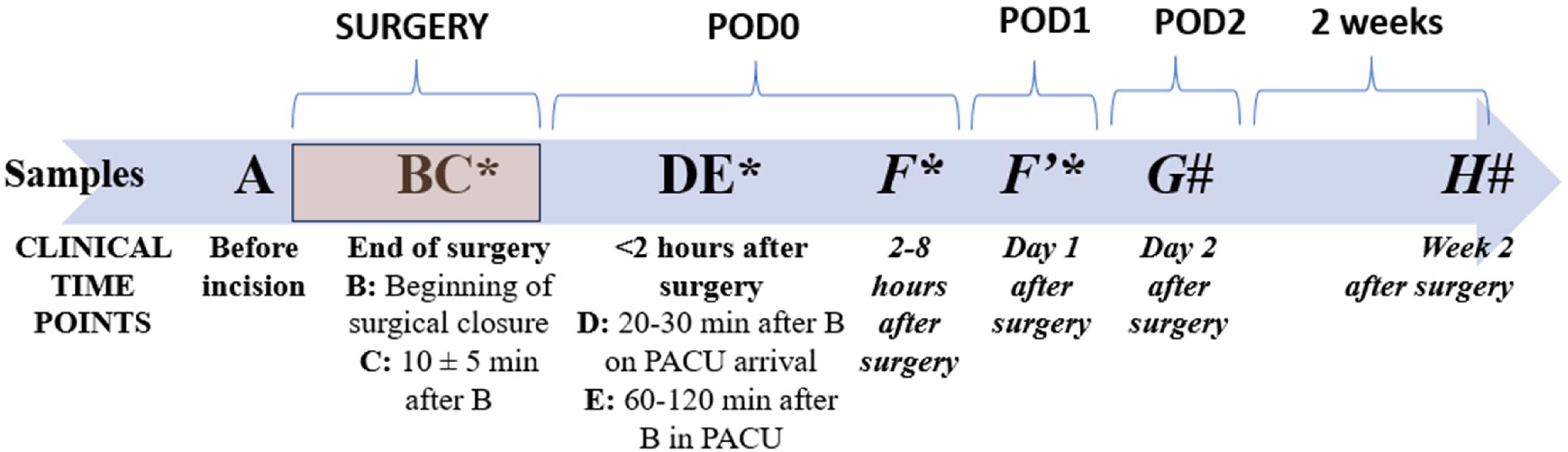
Timeline of blood draws in relation to surgery (marked by rectangle of different shade). Samples are named alphabetically and sequentially. Some samples were combined and averaged based on time of draw (BC and DE). * indicates they were drawn only for spine surgery subjects. # indicates they were drawn only for pectus surgery subjects, due to differences in protocols and to gather more samples. POD: Postoperative day; PACU: Postoperative care unit.

#### 2.2.2 Postoperative pain outcomes

Acute postsurgical pain was defined as a log-transformed compositional outcome of % time spent in moderate – severe pain (NRS>3/10) over POD0-2. Based on area under curve for repeat pain scores measured over POD0-2, % time for which pain scores were>3/10 was calculated. Log-ratio transformation was applied as defined as: f= log (average of % time spent in moderate or severe pain over POD0-2/ average of % time spent in mild pain over POD0-2).

CPSP was defined as any pain (NRS>3/10) beyond 3 months post-surgery. NRS cut-offs of 3/10 depict moderate/severe pain, are associated with functional disability, and have been described as a predictor for the persistence of pain [17].

### 2.3 Cytokine analyses

Cytokine levels were assayed using the Luminex multiplex (Milliplex) immunoassay bead array technology. Kits from R&D Systems (LXSAHM-12) analyzed chemokine CCL22, epidermal growth factor (EGF), fractalkine, IFNγ, IL-1 receptor antagonist (IL-1ra), IL-2, IL-4, IL-6, IL-8, IL-10, IL-13, and TNFα levels; and Millipore Sigma (HSTCMAG28PMX13BK) analyzed granulocyte-macrophage colony-stimulating factor (GM-CSF), IFNγ, IL-1β, IL-2, IL-4, IL-5, IL-6, IL-7, IL-8, IL-10, IL-12 (p70), IL-13 and TNFα levels and run according to the manufacturer’s protocol. See supplemental figures for sensitivity and precision. Concentrations were calculated from standard curves using recombinant proteins and expressed in pg/ml using Bio-Plex Manager 6.2 (Bio-Rad Laboratories, Hercules, CA).

### 2.4 Statistical analyses

Fluorescence intensities were transferred into R (http:www.r-project.org) statistical software for converting into concentration values. Cytokine concentrations were determined from the standard serial dilutions, and standard curves were obtained from the Bayesian hierarchical 5-parameter logistic models created in the R package nCal [18]. Data cleaning and preparation for cytokine data included several steps (weighting, normality check and log transformation, winsorization, ComBat in R package sva for batch effect adjustment and handling of duplicates).

To test if baseline cytokines was associated with acute pain or CPSP, we first conducted an unadjusted analysis to determine if baseline cytokines were associated with acute pain and compared values between CPSP groups using t-tests (Pooled or Satterthwaite based on the test of equality of variances). We assessed the influence of covariates (age, biological sex, race, PPST, surgical duration, surgery type and morphine equivalents per kg (MEQ) or confounders (associated with both the independent variables and the outcomes). We then conducted multiple linear or logistic regression for the log-ratio transformed acute pain and CPSP outcomes, respectively, after eliminating collinear variables, and adjusting for covariates or confounders.

For testing if longitudinal cytokine concentration changes with time were associated with pain outcomes, we computed cytokine AUC for POD0, 1 and 2 (using cytokine concentration and time approximations based on blood draw timings), where cytokine AUC for POD0 was standardized to 24 hours. Cytokine AUCs were log-transformed before analysis. To evaluate the effect of changes in cytokine on acute pain outcome, mixed effects models with the acute pain as the dependent and log transformed cytokine AUC on POD0-2 as the independent variables were used. To evaluate the association between cytokine and CPSP, mixed effects models with log-transformed cytokine AUC on POD0-2 as the outcome and binary CPSP as the independent variable were used. POD, sex, race, surgical duration, surgery type, and total MEQ were included as covariates and subjected to a random effect. We assessed for collinearity (or extreme confounding) for those variables associated with the outcomes based on tolerance, variance inflation factor (VIF) or generalized variance inflation factor (GVIF) for categorical variables.

We eliminated correlated variables in both multiple regression and mixed effects models. Results from full models (supplementary material) and reduced models, where only covariates with *p* ≤ 0.05 were retained, are presented.

## 3. Results

In all, 112 subjects were included in the study. Patient, surgical and outcome summary statistics are provided in **Table 1**. The majority of subjects were White (97.3%) and female (54.5%) and underwent pectus or spine surgeries under standardized anesthesia/pain protocols. The incidence of CPSP in this cohort was 43.9%, with loss of retention in five subjects. One of the cytokines, EGF, was excluded from the regression analysis because the reported concentrations had the same value across all time points.

**Table 1:** Characteristics of study participants (n=112) and variables.

| Variable | Category | Mean (SD) or Median (IQR) (n=112) | Min, Max | Missing (%) |
| --- | --- | --- | --- | --- |
| Age |  | 15.3 (13.5, 17.0) [n=112] | 9.2, 36.4 | 0 (0.0) |
| Sex | Female | 61 (54.5) |  | 0 (0.0) |
|  | Male | 51 (45.5) |  |  |
| Race | Black | 3 (2.7) |  | 0 (0.0) |
|  | White | 109 (97.3) |  |  |
| Surgical duration (minutes) |  | 147.5 (90.5, 224.0) [n=112] | 58.0, 383.0 | 0 (0.0) |
| Surgery | pectus | 67 (59.8) |  | 0 (0.0) |
|  | spine | 45 (40.2) |  |  |
| PPST |  | 1.0 (0.0, 3.0) [n=77] | 0.0, 9.0 | 35 (31.3) |
| MEQ POD0-2 |  | 1.0 (0.7, 1.4) [n=112] | 0.1, 4.1 | 0 (0.0) |
| CPSP | No | 60 (56.1) |  | 5 (4.5) |
|  | Yes | 47 (43.9) |  |  |
| Acute pain POD0-2 |  | 0.1 (-0.9, 1.2) [n=104] | -8.5, 8.5 | 8 (7.1) |

Note: Acute pain is presented as a: log transformed percentage time in moderate-severe pain : log(average % Time in mod/severe pain on POD0-2 / average %Time in mild pain POD0-2)
PPST: Pediatric Pain Screening Tool
MEQ: Morphine equivalents/kg; POD: Postoperative day
CPSP: Chronic postsurgical pain

A total of 4230 cytokine concentration measures were initially included in the analyses. After data cleaning, the resulting number of observations was 3334. Details of data cleaning, tables of winsorized data, plots of before and after ComBat adjustment (**Supplementary** Fig 1) and cytokine data by CPSP over regrouped time points (**Supplementary** Fig 2) are presented in the supplementary materials. Finally, n=3249 concentrations from subjects who self-identified as belonging to White and Black race were included for association analyses due to the small number of subjects from other races.

### 3.1 Univariate associations between baseline cytokines with postoperative pain outcomes

Using unadjusted model, a series of simple linear regressions showed higher acute pain associated with GM-CSF (β=0.90, SE 0.33, *p*=.008), IL-2 (β=0.79, SE 0.35, *p*=.03), and IL-12 p70 (β=0.92, SE 0.43, *p*=.04) concentrations at baseline (**Table S1)**. Comparison of baseline cytokines between CPSP and controls showed differences only for CCL22 (CPSP 5.90±0.52 pg/ml vs. control 6.51 ± 0.40); *p*=.004) (**Table S2**).

### 3.2 Multivariate adjusted analyses: Baseline cytokines and pain outcomes

Biological sex, and surgery type were associated with CPSP, while MEQ was associated with acute pain outcomes (*p*<.05). Participant’s sex was also associated with IL-2 and MEQ with IL-8 levels (acted as confounders). Thus, in models adjusted for sex, MEQ, and surgery type, a series of analyses were conducted to evaluate the association of baseline cytokines with CPSP and acute pain (**Table S3**). Baseline cytokine concentrations were associated with acute pain for GM-CSF (β=0.95, SE 0.31; *p*=.003), IL-1β (β=0.84, SE 0.36; *p*=.02), IL-2 (β=0.78, SE 0.34; *p*=.03), and IL-12 p70 (β=0.88, SE 0.40; *p*=.03), after controlling for relevant covariates and confounders (**Table 2**). In general, greater cytokine concentration levels were associated with with higher MEQ (*p*<.001). After controlling for relevant covariates and confounders, baseline cytokine concentrations were associated with CPSP for IL-8 (β= -0.39, SE 0.17; *p*=.02) (**Table 3**). Baseline concentrations were associated with increased odds for CPSP in female (*vs*. male) sex (IL-2, IL-4, IL-5, IL-6, IL-8, IL-10, and TNFα) and pectus (vs. spine) surgery patients (IL-8, IL-10).

**Table 2:**
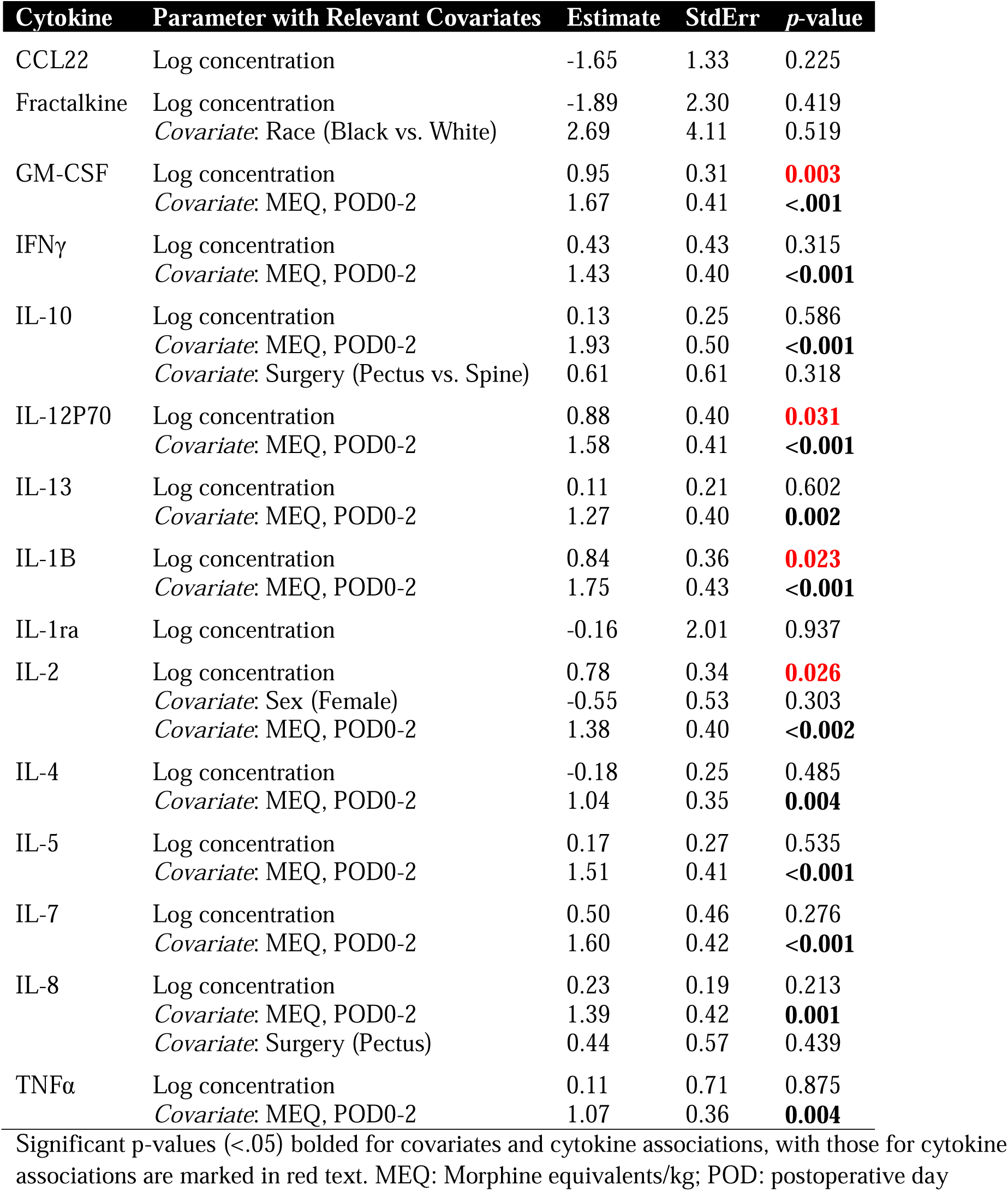
Regression analysis of cytokine log concentration levels at baseline and acute pain (log transformed pain AUC over POD0-1)

**Table 3:** Regression analysis of baseline cytokine concentrations with risk (Odds) of developing chronic postsurgical pain (CPSP)

| **Cytokine** | **Parameter with relevant covariates** | **Odds Ratio (95% CI)** | **Estimate**<br>**$\beta$** | **Std**<br>**Error** | **P -value** |
| --- | --- | --- | --- | --- | --- |
| CCL22 | Log concentration | 0.20 (0.03 - 1.47) | -1.62 | 1.02 | 0.114 |
| Fractalkine | Log concentration | 0.89 (0.09 - 8.71) | -0.12 | 1.17 | 0.918 |
| GM-CSF | Log concentration | 1.35 (0.78 - 2.36) | 0.30 | 0.28 | 0.285 |
| GM-CSF | Covariate: Sex (Female vs. Male) | 2.25 (1.00 - 5.09) | 0.41 | 0.21 | 0.051 |
| IFNγ | Log concentration | 1.29 (0.59 - 2.81) | 0.25 | 0.40 | 0.529 |
| IFNγ | Covariate: Sex (Female vs. Male) | 2.19 (0.97 - 4.94) | 0.39 | 0.21 | 0.059 |
| IL-10 | Log concentration | 0.78 (0.50 - 1.20) | -0.25 | 0.22 | 0.259 |
| IL-10 | Covariate: Sex (Female vs. Male) | 4.27 (1.49 - 12.26) | 0.73 | 0.27 | **0.007** |
| IL-10 | Covariate: Surgery (pectus vs spine) | 3.55 (1.15 - 10.91) | 0.63 | 0.29 | **0.027** |
| IL-12P70 | Log concentration | 1.07 (0.54 - 2.15) | 0.07 | 0.35 | 0.840 |
| IL-12P70 | Covariate: Sex (Female vs. Male) | 2.23 (0.98 - 5.07) | 0.40 | 0.21 | 0.055 |
| IL-13 | Log concentration | 0.79 (0.56 - 1.10) | -0.24 | 0.17 | 0.158 |
| IL-13 | Covariate: Sex (Female vs. Male) | 2.35 (1.03 - 5.36) | 0.43 | 0.21 | **0.042** |
| IL-1B | Log concentration | 1.29 (0.72 - 2.30) | 0.25 | 0.30 | 0.396 |
| IL-1ra | Log concentration | 0.48 (0.05 - 4.65) | -0.74 | 1.16 | 0.525 |
| IL-2 | Log concentration | 1.23 (0.70 - 2.17) | 0.21 | 0.29 | 0.468 |
| IL-2 | Covariate: Sex (Female vs. Male) | 2.46 (1.07 - 5.66) | 0.45 | 0.21 | **0.034** |
| IL-4 | Log concentration | 0.91 (0.59 - 1.39) | -0.10 | 0.22 | 0.646 |
| IL-4 | Covariate: Sex (Female vs. Male) | 2.63 (1.17 - 5.92) | 0.48 | 0.21 | **0.020** |
| IL-5 | Log concentration | 0.84 (0.54 - 1.30) | -0.17 | 0.22 | 0.438 |
| IL-5 | Covariate: Sex (Female vs. Male) | 2.43 (1.07 - 5.55) | 0.44 | 0.21 | **0.035** |
| IL-6 | Log concentration | 0.82 (0.63 - 1.06) | -0.20 | 0.13 | 0.132 |
| IL-6 | Covariate: Sex (Female vs. Male) | 3.09 (1.16 - 8.23) | 0.56 | 0.25 | **0.024** |
| IL-7 | Log concentration | 1.20 (0.54 - 2.67) | 0.18 | 0.41 | 0.655 |
| IL-7 | Covariate: Sex (Female vs. Male) | 2.19 (0.96 - 4.99) | 0.39 | 0.21 | 0.064 |
| IL-8 | Log concentration | 0.68 (0.48 - 0.95) | -0.39 | 0.17 | **0.023** |
| IL-8 | Covariate: Sex (Female vs. Male) | 6.13 (1.98 - 18.92) | 0.91 | 0.29 | **0.002** |
| IL-8 | Covariate: Surgery (pectus vs spine) | 5.53 (1.55 - 19.71) | 0.86 | 0.32 | **0.009** |
| IL-8 | Covariate: MEQ POD0-2 | 1.20 (0.58 - 2.45) | 0.18 | 0.37 | 0.627 |
| TNFα | Log concentration | 0.66 (0.20 - 2.25) | -0.41 | 0.62 | 0.510 |
| TNFα | Covariate: Sex (Female vs. Male) | 2.45 (1.10 - 5.48) | 0.45 | 0.21 | **0.029** |

Significant p-values (<.05) bolded for covariates and cytokine associations, with those for cytokine – pain associations marked in red text. MEQ: Morphine equivalents/kg; POD: postoperative day

### 3.2 Mixed effects model: Association of cytokine concentrations over time with acute pain outcome

Mixed effects models were also adjusted for surgical duration/type, MEQ, sex, and race. In the full model, log cytokine AUC was associated with acute pain in GM-CSF (β=1.38, SE 0.57; *p*=.03), IFNγ (β=1.36, SE 0.6; *p*=.03), IL-1β (β=1.25, SE 0.59; *p*=.03), IL-7 (β=1.65, SE 0.7; *p*=.02), and IL-12 p70 (β=1.17, SE 0.58; *p*=.04) (**Table 4**). POD, surgery type/duration and MEQ were adjusted in several cytokine-acute pain association models (**Table S4**). The final reduced multiple regression model revealed associations with GM-CSF, IFNγ, IL-1β, IL-5, IL-7, and IL-12 p70 (*p*<.05) after adjusting for surgical duration (**Table 4**). Estimates for an association of covariates were positive except for POD2, indicating decreasing acute pain over POD2.

**Table 4.**
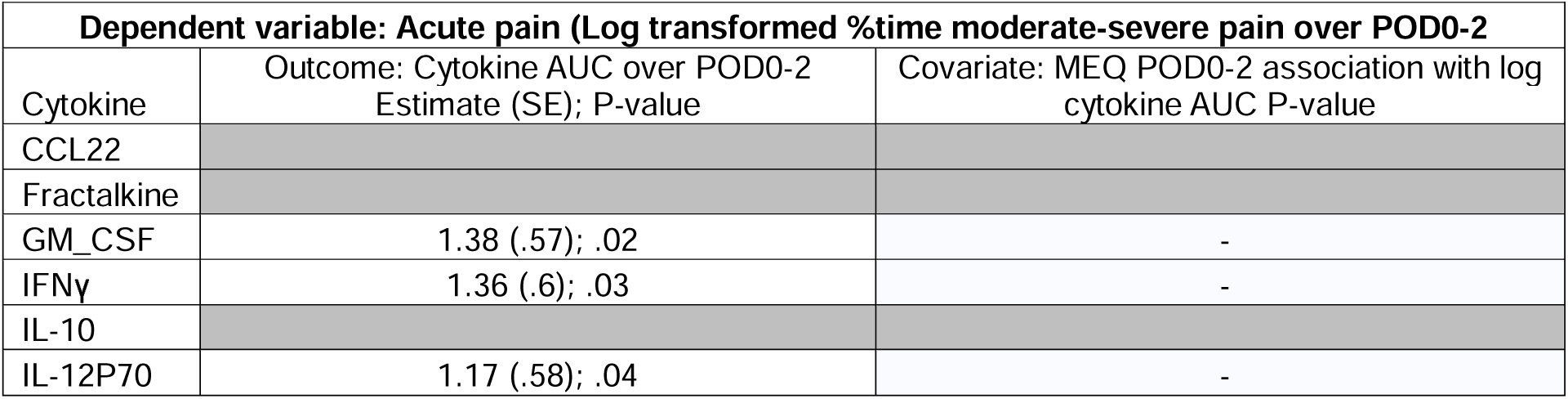
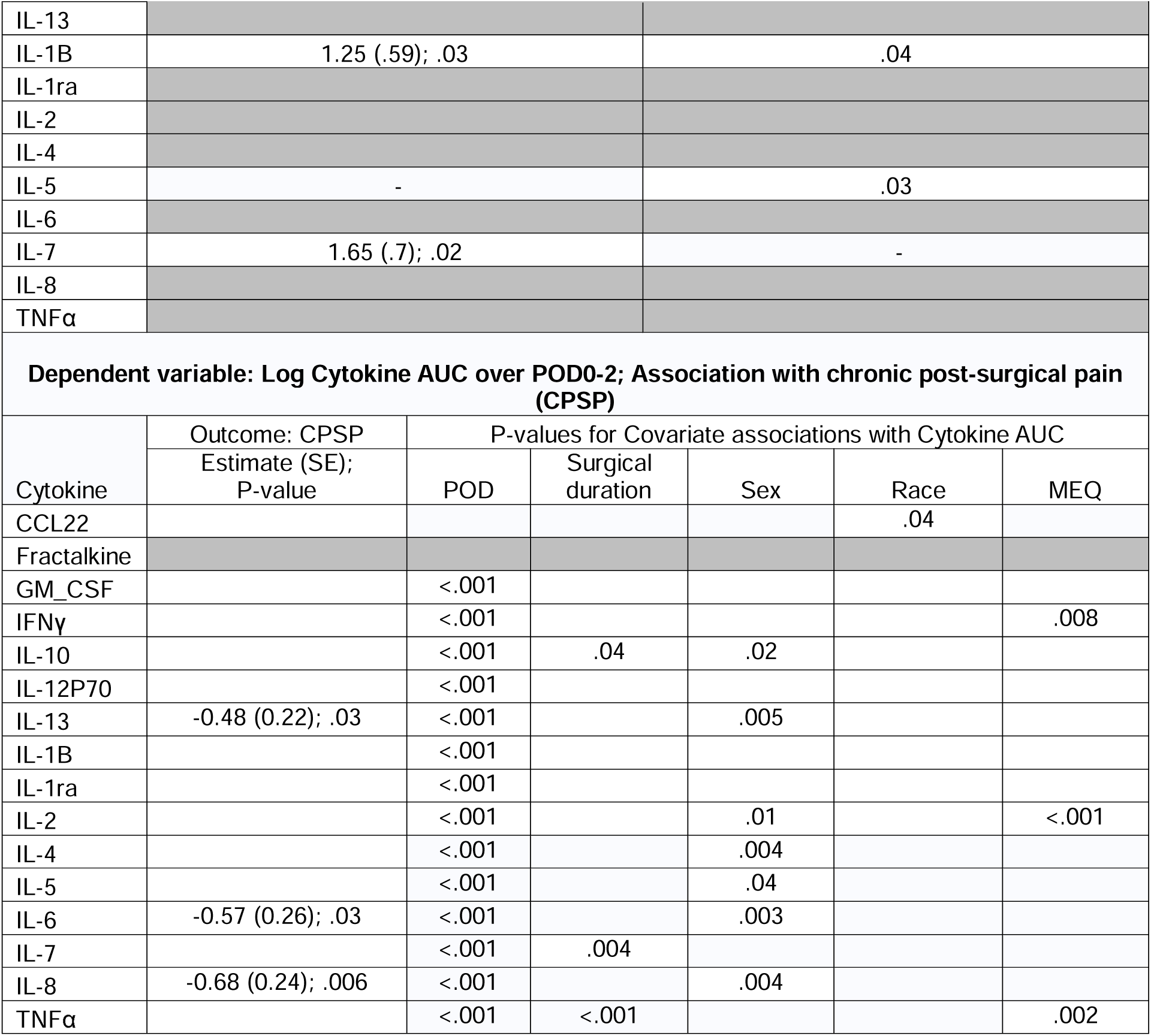
Mixed effects model (reduced model) results for significant associations of pain outcomes and covariates with cytokine concentrations over POD0-2, adjusted for surgical duration.

### 3.4 Mixed effects model: Association of longitudinal cytokine concentrations and CPSP

Line chart of cytokine concentrations over time are provided in **Fig S2**, and the full model is in **Table S5**. CPSP was significantly associated with log cytokine AUC for IL-6 (β= - 0.57, SE 0.26; *p*=.03), IL-8 (β= -0.68, SE 0.24; *p*=.006), and IL-13 (β= -0.48, SE 0.22; *p*=.03) (**Table 4**). Postoperative day and surgical duration were adjusted in the reduced model. Estimates for association with log cytokine AUC were negative for pectus surgery and Black race and positive for POD (POD1 and POD2) and MEQ. (**Table S5**)

## 4. Discussion

In this prospective study of adolescents undergoing pectus or spine deformity correction surgery, cytokines were assayed from blood samples collected preoperatively and over six time points over POD0-2 and week 2 after surgery. The aims were to determine associations of baseline and longitudinal cytokine changes over POD0-2 with acute and chronic postoperative pain, adjusted for other covariates. We identified POD, sex, MEQ and surgery type/duration as covariates, as these were associated with cytokine and pain outcomes. Multivariate regression models revealed that higher baseline (i.e., preoperative) levels of GM-CSF, IL-1β, IL-2, and IL-12 p70 concentrations were associated with acute pain (i.e., % time in moderate-severe pain after surgery) while higher levels of baseline IL-8 decreased the odds for CPSP. Mixed effects models demonstrated that longitudinal postoperative cytokine concentrations of IL-6, IL-8, and IL-13 were protective for CPSP (i.e. increased concentration decreased odds of developing CPSP), while higher longitudinal postoperative concentrations of GM-CSF, IFNγ, IL-1β, IL-7, and IL-12 p70 were associated with higher acute pain. Collectively, our data identified distinct cytokine signatures that are associated with differential outcomes for acute and chronic postsurgical pain. Severity of acute pain was predicted by elevations of mostly pro-inflammatory, chemotactic, and proliferative cytokines, while likelihood of developing chronic pain was associated with baseline and postoperative elevations of IL-6, IL-8, and IL-13, which suggest a pain-resolving mechanism associated with these immune mediators.

The immediate postoperative period is characterized by an orchestrated release of cytokines in response to surgical trauma. We found that baseline or overall AUC of GM-CSF, IFNγ, IL-1β, IL-2, IL-7, and IL-12 p70 were higher with % time in moderate-severe pain on POD0-2. Several of these cytokines are known to be pro-inflammatory; some are secreted by T helper cells and lead to the growth/differentiation of monocytes (GM-CSF) and proliferation of B cells, T cells, and natural killer (NK) cells (IL-2, IL-7). In contrast, others are released by macrophages and act on NK cells (IL-12 p70) and lead to IFNγ release, which activates macrophages, neutrophils and NK cells and promotes cell-mediated immunity. IL-6 and TNFα have also been implicated in sensitizing pain pathways and amplifying nociceptive signals[7]. However, our study did not find them to be associated with pain outcomes. This aligns with other studies in adults. After hip arthroplasty serial IL-6 concentrations on POD1 were found to predict disability but not pain on discharge [19]., or long-term disability at one and six months after surgery. Another study evaluating preoperative IL-6 and TNFα did not predict early pain or function after total hip arthroplasty [20]. These conflicting reports suggest that individual cytokine concentrations are not uniformly predictive of pain outcomes, and further work is required to uncover specific determinants of inflammatory responses in the postoperative period.

Traditionally, cytokines such as IL-1 receptor antagonist, IL-4, IL-10, IL-11, and IL-13 are considered anti-inflammatory. Low blood levels of IL-4 and IL-10 were reported to be associated with chronic widespread pain [21]. However, we did not find associations between IL-4 or IL-10 with CPSP. We did find IL-13 to be protective for CPSP. IL-13 (and IL-4) belong to the T helper 2 (Th2) cytokine family and have been shown to play a role in downmodulating inflammatory responses in rheumatoid arthritis, induce macrophage polarization from a pro-inflammatory M1 to an anti-inflammatory M2 phenotype [22]. Studies in mice have shown that IL-13 released from T cells can polarize macrophages into an M2-like phenotype, and the resulting production of anti-inflammatory cytokines from these polarized macrophages was found to be critical for the resolution of chemotherapy-induced peripheral neuropathy.[23]. IL-13 signaling uses the JAK-signal transducer and activator of the transcription (STAT) pathway, specifically STAT6 [24], and these pathways may be future intervention targets. We also found baseline IL-8 and postoperative IL-8 AUC over POD0-2 to be protective for CPSP. However, this is not aligned with its known pro-inflammatory role and its association with pain in other studies [25]. Instead, our data support the notion that IL-8 may promote the recruitment or expansion of endogenous pain-resolving immune cells [add citations]. In support of this, patients with transcriptional responses associated with acute neutrophil activation after injury were less likely to transition from acute to chronic lower back pain[26]. Furthermore, female sex remained a significant factor associated with higher CPSP in these models. Our findings are aligned with those of a study of inflammatory markers in chronic pelvic pain patients. They reported IL-8 was protective with a sex-specific effect: women with pelvic pain showed a negative relationship between IL-8 and widespread pain, while men did not [27]. Female sex was also a covariate for AUC cytokine associations with CPSP for IL-4, IL-6, IL-8, and IL-13 cytokines. In an experimental study of sex effects on lipopolysaccharide (LPS) induced immune activation, visceral and musculoskeletal hyperalgesia was found to occur in both men and women, irrespective of biological sex. However, the LPS-induced cytokine increases in women were more pronounced [28]. Thus, future studies should investigate sex-cytokine interactions in pain as a biological mechanism for sex differences in pain.

We identified postsurgical time-dependent changes in cytokine levels with increased cytokine levels on POD1 followed by a decline on POD2. This is similar to the results of another study in 11 patients which found that five cytokines (IL-4, IL-6, IL-10, IL-12 p70 and IL-13) in plasma and ten cytokines in the cerebrospinal fluid (CSF) (IFNγ, IL-1β, IL-2, IL-4, IL-6, IL-8, IL-10, IL-12 p70, IL-13, and TNFα) rose significantly following femoral neck surgery [29]. However, the study did not report associations with clinical pain. Interestingly, neuroinflammatory responses (in CSF) were not shown to correlate with plasma levels. This is surprising as postsurgical studies in elderly patients found plasma TNFα, IL-6, and IL-10 levels to be independent predictors for adverse central postoperative outcomes such as cognitive deficits [30]. Future studies need to understand better the correlation of neuroinflammatory markers in clinical pain cohorts.

Surgery type (pectus vs spine) was associated with IL-8 and IL-10 at baseline in CPSP and acute pain association models. Prior research has shown similar differences with IL-6 increases with more major surgery (aortic vs. hernia surgery) and in those with complications (N=3) [31] Opioid consumption was identified as a covariate for the association of baseline cytokine as well as cytokine exposure (AUC) for several cytokines (GM-CSF, IL-1β, IL2-8, IL-10, IL-12 p70, IL-13, TNFα) with acute pain, even when surgery type and duration were adjusted for in the models.

Our findings support the hypothesis that pro-inflammatory antagonists or anti-inflammatory agents could attenuate acute and chronic postsurgical pain. Our results predict that pharmacological or immunomodulatory approaches to broadly inhibit circulating pro-inflammatory cytokines would improve outcomes associated with acute pain, while interventions to elevate IL-6, IL-8, and IL-13 in the immediate postoperative period could prevent the transition from acute to chronic pain. Although it remains an ongoing challenge to suppress pain-promoting inflammation while promoting pain-resolving immune responses, there is evidence that immunotherapies may improve the treatment of ongoing pain[32, 33] . For example, anti-TNFα agents such as etanercept have found promise in inflammatory conditions such as rheumatoid arthritis [34]. Recently, IL-13 administration in mice was found to reverse inflammatory macrophage-dependent neuropathic pain *via* a phenotype shift toward suppressive macrophages [35]. Our results add to the evolving understanding of cytokine involvement in postoperative pain, supporting emerging strategies targeting cytokine signaling to alleviate postoperative pain in children. Of note, multimodal analgesia components such as intravenous lidocaine and dexamethasone have been investigated for their anti-inflammatory effects and potential analgesic benefits after major musculoskeletal surgeries such as spine fusion, with mixed evidence regarding their benefits [36, 37].

### 4.1 Limitations

One limitation of our study is that we did not adjust statistical significance for multiple tests. Despite this, to our knowledge, this might be the first study of longitudinal cytokines in relation to acute and chronic post-surgical pain in a well characterized, prospective, pediatric cohort. Moreover, the generalization over two surgeries makes our findings generalizable. Also, although the data was analyzed in different batches, we used high rigor in normalizing and analyzing the cytokine data. Despite the study’s observational nature, spine and pectus surgical patients received standardized protocols, minimizing differences in pain management regimens.

### 4.2 Conclusions

In conclusion, this paper describes the association between cytokines and acute and chronic postoperative pain in pediatric patients. Cytokine measurements perioperatively may not only serve as biomarkers and enable prediction of pain-related risk but also suggest potential tailored interventions, thus paving the way for more personalized and effective pain management in the pediatric surgical setting. Future studies need to elucidate sex interactions with immune signaling and surgery-specific differences.

## Data Availability

All data produced in the present study are available upon reasonable request to the authors

## Acknowledgments

The authors gratefully acknowledge Maria E Ashton, MS, RPh, MBA, Medical Writer, Department of Anesthesiology, Cincinnati Children’s Hospital Medical Center, for providing writing assistance, editing, and proofreading.

## References

[1] Lee CS, Merchant S, Chidambaran V. Postoperative Pain Management in Pediatric Spinal Fusion Surgery for Idiopathic Scoliosis. Paediatr Drugs. 2020;22:575–601.

[2] Rabbitts JA, Fisher E, Rosenbloom BN, Palermo TM. Prevalence and Predictors of Chronic Postsurgical Pain in Children: A Systematic Review and Meta-Analysis. J Pain. 2017;18:605–14.

[3] Chidambaran V, Ding L, Moore DL, Spruance K, Cudilo EM, Pilipenko V, et al. Predicting the pain continuum after adolescent idiopathic scoliosis surgery: A prospective cohort study. Eur J Pain. 2017;21:1252–65.

[4] Weledji EP. The role of cytokines in enhanced recovery after surgery. IJS Short Reports. 2021;6.

5. Hsing C-H, Wang J-J. Clinical implication of perioperative inflammatory cytokine alteration. Acta Anaesthesiologica Taiwanica. 2015;53:23–8.

[6] Bosch DJ, Nieuwenhuijs-Moeke GJ, van Meurs M, Abdulahad WH, Struys M. Immune Modulatory Effects of Nonsteroidal Anti-inflammatory Drugs in the Perioperative Period and Their Consequence on Postoperative Outcome. Anesthesiology. 2022;136:843–60.

[7] Grace PM, Hutchinson MR, Maier SF, Watkins LR. Pathological pain and the neuroimmune interface. Nat Rev Immunol. 2014;14:217–31.

[8] Si HB, Yang TM, Zeng Y, Zhou ZK, Pei FX, Lu YR, et al. Correlations between inflammatory cytokines, muscle damage markers and acute postoperative pain following primary total knee arthroplasty. BMC Musculoskelet Disord. 2017;18:265.

[9] Watkins LR, Maier SF, Goehler LE. Immune activation: the role of pro-inflammatory cytokines in inflammation, illness responses and pathological pain states. Pain. 1995;63:289–302.

[10] Gandhi R, Santone D, Takahashi M, Dessouki O, Mahomed NN. Inflammatory predictors of ongoing pain 2 years following knee replacement surgery. Knee. 2013;20:316–8.

[11] Azim S, Nicholson J, Rebecchi MJ, Galbavy W, Feng T, Rizwan S, et al. Interleukin-6 and leptin levels are associated with preoperative pain severity in patients with osteoarthritis but not with acute pain after total knee arthroplasty. Knee. 2018;25:25–33.

[12] Narayanasamy S, Yang F, Ding L, Geisler K, Glynn S, Ganesh A, et al. Pediatric Pain Screening Tool: A Simple 9-Item Questionnaire Predicts Functional and Chronic Postsurgical Pain Outcomes After Major Musculoskeletal Surgeries. J Pain. 2022;23:98–111.

[13] Chidambaran V, Zhang X, Geisler K, Stubbeman BL, Chen X, Weirauch MT, et al. Enrichment of Genomic Pathways Based on Differential DNA Methylation Associated With Chronic Postsurgical Pain and Anxiety in Children: A Prospective, Pilot Study. J Pain. 2019;20:771–85.

[14] Vandenbroucke JP, von Elm E, Altman DG, Gotzsche PC, Mulrow CD, Pocock SJ, et al. Strengthening the Reporting of Observational Studies in Epidemiology (STROBE): explanation and elaboration. Epidemiology. 2007;18:805–35.

[15] von Baeyer CL. Numerical rating scale for self-report of pain intensity in children and adolescents: recent progress and further questions. Eur J Pain. 2009;13:1005–7.

[16] Simons LE, Smith A, Ibagon C, Coakley R, Logan DE, Schechter N, et al. Pediatric Pain Screening Tool: rapid identification of risk in youth with pain complaints. Pain. 2015;156:1511–8.

[17] Gerbershagen HJ, Rothaug J, Kalkman CJ, Meissner W. Determination of moderate-to-severe postoperative pain on the numeric rating scale: a cut-off point analysis applying four different methods. Br J Anaesth. 2011;107:619–26.

[18] Fong Y, Sebestyen K, Yu X, Gilbert P, Self S. nCal: an R package for non-linear calibration. Bioinformatics. 2013;29:2653–4.

[19] Hall GM, Peerbhoy D, Shenkin A, Parker CJ, Salmon P. Relationship of the functional recovery after hip arthroplasty to the neuroendocrine and inflammatory responses. Br J Anaesth. 2001;87:537–42.

[20] Poehling-Monaghan KL, Taunton MJ, Kamath AF, Trousdale RT, Sierra RJ, Pagnano MW. No Correlation Between Serum Markers and Early Functional Outcome After Contemporary THA. Clin Orthop Relat Res. 2017;475:452–62.

[21] Uçeyler N, Valenza R, Stock M, Schedel R, Sprotte G, Sommer C. Reduced levels of antiinflammatory cytokines in patients with chronic widespread pain. Arthritis Rheum. 2006;54:2656–64.

[22] Iwaszko M, Biały S, Bogunia-Kubik K. Significance of Interleukin (IL)-4 and IL-13 in Inflammatory Arthritis. Cells. 2021;10.

[23] Singh SK, Krukowski K, Laumet GO, Weis D, Alexander JF, Heijnen CJ, et al. CD8+ T cell-derived IL-13 increases macrophage IL-10 to resolve neuropathic pain. JCI Insight. 2022;7.

[24] Takeda K, Kamanaka M, Tanaka T, Kishimoto T, Akira S. Impaired IL-13-mediated functions of macrophages in STAT6-deficient mice. J Immunol. 1996;157:3220–2.

[25] Hedderson WC, Borsa PA, Fillingim RB, Coombes SA, Hass CJ, George SZ. Plasma Concentrations of Select Inflammatory Cytokines Predicts Pain Intensity 48 Hours Post-Shoulder Muscle Injury. Clin J Pain. 2020;36:775–81.

[26] Parisien M, Lima LV, Dagostino C, El-Hachem N, Drury GL, Grant AV, et al. Acute inflammatory response via neutrophil activation protects against the development of chronic pain. Sci Transl Med. 2022;14:eabj9954.

[27] Karshikoff B, Martucci KT, Mackey S. Relationship Between Blood Cytokine Levels, Psychological Comorbidity, and Widespreadness of Pain in Chronic Pelvic Pain. Frontiers in Psychiatry. 2021;12.

[28] Wegner A, Elsenbruch S, Rebernik L, Roderigo T, Engelbrecht E, Jäger M, et al. Inflammation-induced pain sensitization in men and women: does sex matter in experimental endotoxemia? Pain. 2015;156:1954–64.

[29] Fertleman M, Pereira C, Dani M, Harris BHL, Di Giovannantonio M, Taylor-Robinson SD. Cytokine changes in cerebrospinal fluid and plasma after emergency orthopaedic surgery. Scientific Reports. 2022;12:2221.

[30] Sun T, Wang X, Liu Z, Chen X, Zhang J. Plasma concentrations of pro- and anti-inflammatory cytokines and outcome prediction in elderly hip fracture patients. Injury. 2011;42:707–13.

[31] Baigrie RJ, Lamont PM, Kwiatkowski D, Dallman MJ, Morris PJ. Systemic cytokine response after major surgery. Br J Surg. 1992;79:757–60.

[32] Kavelaars A, Heijnen CJ. Immune regulation of pain: Friend and foe. Sci Transl Med. 2021;13:eabj7152.

[33] Fiore NT, Debs SR, Hayes JP, Duffy SS, Moalem-Taylor G. Pain-resolving immune mechanisms in neuropathic pain. Nat Rev Neurol. 2023;19:199–220.

[34] Hung AL, Lim M, Doshi TL. Targeting cytokines for treatment of neuropathic pain. Scandinavian Journal of Pain. 2017;17:287–93.

[35] Kiguchi N, Sakaguchi H, Kadowaki Y, Saika F, Fukazawa Y, Matsuzaki S, et al. Peripheral administration of interleukin-13 reverses inflammatory macrophage and tactile allodynia in mice with partial sciatic nerve ligation. Journal of Pharmacological Sciences. 2017;133:53–6.

[36] Dewinter G, Moens P, Fieuws S, Vanaudenaerde B, Van de Velde M, Rex S. Systemic lidocaine fails to improve postoperative morphine consumption, postoperative recovery and quality of life in patients undergoing posterior spinal arthrodesis. A double-blind, randomized, placebo-controlled trial. British Journal of Anaesthesia. 2017;118:576–85.

[37] Farag E, Ghobrial M, Sessler DI, Dalton JE, Liu J, Lee JH, et al. Effect of Perioperative Intravenous Lidocaine Administration on Pain, Opioid Consumption, and Quality of Life after Complex Spine Surgery. Anesthesiology. 2013;119:932–40.

