## Supplemental tables_figures for "The Role of Cytokines in Acute and Chronic Postsurgical Pain in Pediatric Patients after Major Musculoskeletal Surgeries"

**Corresponding author:**

Vidya Chidambaran, MD, MS

**Supplementary Methods**

**Data cleaning and preparation**

1. Read in **4230** row of observations (**118** unique subjects).

### 2. Baseline distribution and normality check

Density and QQ plots as well as Shapiro-Wilk’s normality test followed by Log transformation to reduce the influence of extremely high values and improve normality of the data. However, some cytokine still had skewed distribution. To further reduce the impact of extreme values, winsorization was applied.

3. Winsorization: Replace extreme values for each marker with 25 percentile - 3*IQR or 75 percentile + 3* IQR

Below is the summary of number of observations and number winsorized (n.win) as well as the min, max and upper/lower bounds. The summary shows that winsorization has effects mostly on extremely low values. All EGF values become the same, 1.13, after winsorization.

**|cytokine	|	n| n.chg|	min| max| lowerbd| upperbd|**

|CCL22 | 136| 1| 2.79| 7.16| 3.60| 8.55|

|EGF | 112| 10| 1.82| 4.45| 1.82| 1.82|

|Fractalkine | 135| 10| 5.92| 8.19| 5.93| 8.25|

|GM_CSF | 229| 5| -0.51| 4.62| -0.25| 6.00|

|IFNγ | 282| 0| -1.20| 5.86| -2.44| 7.39|

|IL-10 | 319| 0| -0.36| 5.66| -2.83| 7.12|

|IL-12P70 | 227| 1| -1.61| 3.03| -1.93| 2.57|

|IL-13 | 292| 0| -2.30| 6.67| -10.05| 16.33|

|IL-1B | 227| 0| -1.61| 3.50| -5.51| 6.51|

|IL-1ra | 135| 0| 4.35| 8.59| 3.70| 9.72|

|IL-2 | 316| 0| -1.61| 4.61| -8.81| 10.91|

|IL-4 | 326| 0| -0.11| 7.10| -2.37| 9.53|

|IL-5 | 227| 0| -1.61| 2.86| -3.03| 4.05|

|IL-6 | 348| 0| -2.30| 5.07| -5.00| 9.96|

|IL-7 | 227| 0| 0.88| 5.53| -1.21| 8.65|

|IL-8 | 361| 0| -1.61| 5.76| -4.81| 9.59|

|TNFα | 331| 12| -1.61| 2.84| 0.18| 2.25|

Density and QQ plots as well as Shapiro-Wilk’s normality test results after winsorization were checked.

4. Adjusting for batch effect using ComBat function in R package, sva

ComBat function requires at least two samples in each batch in each row, i.e., at least two non-missing values in each batch for each cytokine. Cytokines, GM_CSF, IL-1B, IL-5, IL-7, and IL-12P70 are only measured in batches 2 and 3 and cytokines, CCL22, EGF, Fractalkine, IL-1ra are only measured in batch 1. Therefore, adjustment was done for cytokines without missing batches and then GM_CSF, IL-1B, IL-5, IL-7, and IL-12P70 in only batches 2 and 3. CCL22, EGF, Fractalkine, IL-1ra were not adjusted.

Plots of batch effect before and after ComBat adjustment can be found in Supplementary Figure 1 (example IL2)

5. Handling duplicates
   a. A total of 271 observations contain two duplicated copies by visit by cytokine (the rest are unique observations).
   b. The following steps were used to handle the duplicates:

1. If one Data_Lab value is NA, use the cytokine value and weight in the other observation (n=221)

2. If both Data_Lab values are non-NA,

a. If weight is the same, use average of both observations (n=25)

b. If one weight is 0.5 and the other is 1, use the cytokine value from observation with weight of 1 (n=1)

3. If both Data_Lab values are NA, assign NA cytokine value and weight (n=24)

c. After removing duplicated pairs, the resulting number of observations is **3959**.
6. Regrouping:
   a. A
   b. B, C as BC
   c. D, E as DE
   d. The rest as is

After regrouping, the resulting number of observations is **3334**, including 580 observations with weight 0.5 and 2735 observations with weight 1.

Due to small number of other races (n=6), only “white” and “black” were retained - number of observations reduce to **3249**.

Table S1: Acute pain (%time in mod-severe pain) with baseline cytokine

|  | Weighted linear regression | | | Weighted Spearman’s correlation | |
| --- | --- | --- | --- | --- | --- |
| Cytokine | Estimate | SE | P-Value | Correlation Coefficient | P-Value |
| CCL22 | -1.65 | 1.33 | 0.225 | -0.25 | 0.225 |
| Fractalkine | -1.33 | 2.11 | 0.534 | -0.08 | 0.695 |
| GM_CSF | 0.90 | 0.33 | 0.008 | 0.23 | 0.022 |
| IFNγ | 0.45 | 0.45 | 0.320 | 0.10 | 0.319 |
| IL-10 | 0.03 | 0.25 | 0.892 | 0.00 | 0.988 |
| IL-12P70 | 0.92 | 0.43 | 0.035 | 0.22 | 0.031 |
| IL-13 | 0.08 | 0.22 | 0.726 | 0.04 | 0.685 |
| IL-1B | 0.79 | 0.39 | 0.046 | 0.20 | 0.056 |
| IL-1ra | -0.16 | 2.01 | 0.937 | -0.02 | 0.937 |
| IL-2 | 0.79 | 0.35 | 0.028 | 0.24 | 0.021 |
| IL-4 | -0.11 | 0.26 | 0.669 | -0.04 | 0.667 |
| IL-5 | 0.13 | 0.29 | 0.651 | 0.04 | 0.718 |
| IL-6 | -0.07 | 0.14 | 0.626 | -0.04 | 0.717 |
| IL-7 | 0.52 | 0.49 | 0.292 | 0.11 | 0.292 |
| IL-8 | 0.11 | 0.18 | 0.539 | 0.05 | 0.643 |
| TNFα | -0.10 | 0.73 | 0.889 | -0.03 | 0.770 |

**Table S2. Comparing baseline cytokines between CPSP 0 vs. 1 using two sample t-tests with weights (n total obs = 1510).**

|  | **CPSP_Yes** | | **CPSP_No** | |  |  |
| --- | --- | --- | --- | --- | --- | --- |
| **Cytokine** | **n** | **Mean (SD)** | **n** | **Mean (SD)** | **p-value** | **Variance** |
| CCL22 | 8 | 5.90 (0.52) | 17 | 6.51 (0.40) | 0.004 | Equal |
| EGF | 4 | 1.82 (0.00) | 15 | 1.82 (0.00) | NA | NA |
| Fractalkine | 8 | 7.04 (0.15) | 17 | 7.11 (0.40) | 0.589 | Unequal |
| GM_CSF | 25 | 3.01 (0.71) | 70 | 2.83 (0.74) | 0.294 | Equal |
| IFNγ | 25 | 2.63 (0.69) | 70 | 2.48 (0.47) | 0.327 | Unequal |
| IL-10 | 25 | 1.92 (0.88) | 68 | 1.95 (0.99) | 0.865 | Equal |
| IL-12P70 | 25 | 0.44 (0.64) | 68 | 0.43 (0.48) | 0.943 | Equal |
| IL-13 | 26 | 2.75 (1.53) | 69 | 3.04 (1.01) | 0.390 | Unequal |
| IL-1B | 25 | 0.45 (0.81) | 69 | 0.31 (0.58) | 0.491 | Unequal |
| IL-1ra | 8 | 5.98 (0.29) | 17 | 6.08 (0.41) | 0.517 | Equal |
| IL-2 | 25 | 1.50 (0.76) | 67 | 1.22 (0.68) | 0.119 | Equal |
| IL-4 | 26 | 3.65 (0.96) | 72 | 3.72 (0.92) | 0.737 | Equal |
| IL-5 | 25 | 0.57 (0.82) | 69 | 0.61 (0.82) | 0.854 | Equal |
| IL-6 | 23 | 2.00 (1.52) | 65 | 2.23 (1.45) | 0.545 | Equal |
| IL-7 | 25 | 3.76 (0.61) | 68 | 3.60 (0.49) | 0.198 | Equal |
| IL-8 | 27 | 1.86 (1.28) | 72 | 2.14 (1.32) | 0.369 | Equal |
| TNFα | 26 | 1.13 (0.31) | 71 | 1.07 (0.29) | 0.413 | Equal |

Table S3: Model summary (Multiple regression for baseline cytokine with pain outcomes)

| Cytokine | CPSP | transformed %AUC POD0-2 |
| --- | --- | --- |
| CCL22 | concentration_bl | concentration_bl |
| Fractalkine | concentration_bl | concentration_bl + race |
| GM_CSF | concentration_bl + sex | concentration_bl + MEQ POD0-2 |
| IFNγ | concentration_bl + sex | concentration_bl + MEQ POD0-2 |
| IL-10 | concentration_bl + sex + surgery type | concentration_bl + MEQ POD0-2 + surgery_type |
| IL-12P70 | concentration_bl + sex | concentration_bl + MEQ POD0-2 |
| IL-13 | concentration_bl + sex | concentration_bl + MEQ POD0-2 |
| IL-1B | concentration_bl | concentration_bl + MEQ POD0-2 |
| IL-1ra | concentration_bl | concentration_bl |
| IL-2 | concentration_bl + sex | concentration_bl + sex + MEQ POD0-2 |
| IL-4 | concentration_bl + sex | concentration_bl + MEQ POD0-2 |
| IL-5 | concentration_bl + sex | concentration_bl + MEQ POD0-2 |
| IL-6 | concentration_bl + sex | concentration_bl + MEQ POD0-2 |
| IL-7 | concentration_bl + sex | concentration_bl + MEQ POD0-2 |
| IL-8 | concentration_bl + sex + surgery type + MEQ | concentration_bl + MEQ POD0-2 + surgery_type |
| TNFα | concentration_bl + sex | concentration_bl + MEQ POD0-2 |

Table S4. Mixed effects full model: acute pain (dependent variable) vs log cytokine area under curve (longitudinal changes in cytokine concentrations over postoperative days 0-2 (POD 0-2)

| **Cytokine** | **Parameters/Covariates** | **level** | **Type3_Pvalue** | **Estimate** | **StdErr** |
| --- | --- | --- | --- | --- | --- |
| CCL22 | log_cytokine_AUC |  | 0.666 | -0.65 | 1.48 |
| CCL22 | POD | POD1 | 0.240 | 2.51 | 2.03 |
| CCL22 | sex | Male | 0.361 | -2.69 | 2.85 |
| CCL22 | race | Black | 0.259 | 6.40 | 5.51 |
| CCL22 | surgical duration |  | 0.405 | 0.01 | 0.01 |
| CCL22 | surgical type |  | . | . | . |
| CCL22 | MEQ POD0-2 |  | 0.619 | 0.78 | 1.55 |
| EGF | log_cytokine_AUC |  | 0.295 | 3.01 | 2.82 |
| EGF | POD | POD1 | 0.470 | 1.76 | 2.37 |
| EGF | sex | Male | 0.367 | -2.46 | 2.64 |
| EGF | race |  | . | . | . |
| EGF | surgical duration |  | 0.585 | 0.01 | 0.01 |
| EGF | surgical type |  | . | . | . |
| EGF | MEQ POD0-2 |  | 0.243 | 2.32 | 1.94 |
| Fractalkine | log_cytokine_AUC |  | 0.531 | 1.17 | 1.84 |
| Fractalkine | POD | POD1 | 0.433 | 1.58 | 1.97 |
| Fractalkine | sex | Male | 0.309 | -3.02 | 2.88 |
| Fractalkine | race | Black | 0.104 | 8.99 | 5.30 |
| Fractalkine | surgical duration |  | 0.538 | 0.01 | 0.01 |
| Fractalkine | surgical type |  | . | . | . |
| Fractalkine | MEQ POD0-2 |  | 0.612 | 0.79 | 1.55 |
| GM_CSF | log_cytokine_AUC |  | 0.005 | 1.59 | 0.56 |
| GM_CSF | POD | POD1 | 0.028 | 4.48 | 2.45 |
| GM_CSF | POD | POD2 | 0.028 | -2.02 | 1.40 |
| GM_CSF | sex | Male | 0.960 | 0.05 | 0.97 |
| GM_CSF | race | Black | 0.371 | 2.14 | 2.38 |
| GM_CSF | surgical duration |  | 0.951 | 0.0004 | 0.01 |
| GM_CSF | surgical type | pectus | 0.036 | 3.80 | 1.79 |
| GM_CSF | MEQ POD0-2 |  | 0.007 | 4.25 | 1.55 |
| IFNγ | log_cytokine_AUC |  | 0.012 | 1.51 | 0.60 |
| IFNγ | POD | POD1 | 0.087 | 2.18 | 2.09 |
| IFNγ | POD | POD2 | 0.087 | -2.35 | 1.49 |
| IFNγ | sex | Male | 0.697 | 0.38 | 0.98 |
| IFNγ | race | Black | 0.242 | 2.87 | 2.41 |
| IFNγ | surgical duration |  | 0.809 | 0.002 | 0.01 |
| IFNγ | surgical type | pectus | 0.070 | 3.16 | 1.72 |
| IFNγ | MEQ POD0-2 |  | 0.019 | 3.38 | 1.42 |
| IL-10 | log_cytokine_AUC |  | 0.679 | -0.17 | 0.40 |
| IL-10 | POD | POD1 | 0.462 | 2.31 | 1.89 |
| IL-10 | POD | POD2 | 0.462 | -0.06 | 1.31 |
| IL-10 | sex | Male | 0.854 | -0.18 | 0.99 |
| IL-10 | race | Black | 0.131 | 3.88 | 2.51 |
| IL-10 | surgical duration |  | 0.926 | 0.001 | 0.01 |
| IL-10 | surgical type | pectus | 0.379 | 1.54 | 1.74 |
| IL-10 | MEQ POD0-2 |  | 0.039 | 2.53 | 1.21 |
| IL-12P70 | log_cytokine_AUC |  | 0.016 | 1.45 | 0.59 |
| IL-12P70 | POD | POD1 | 0.025 | 5.14 | 2.44 |
| IL-12P70 | POD | POD2 | 0.025 | -1.77 | 1.42 |
| IL-12P70 | sex | Male | 0.769 | 0.29 | 0.99 |
| IL-12P70 | race | Black | 0.245 | 2.77 | 2.38 |
| IL-12P70 | surgical duration |  | 0.768 | 0.002 | 0.01 |
| IL-12P70 | surgical type | pectus | 0.036 | 3.86 | 1.82 |
| IL-12P70 | MEQ POD0-2 |  | 0.006 | 4.35 | 1.56 |
| IL-13 | log_cytokine_AUC |  | 0.837 | -0.08 | 0.38 |
| IL-13 | POD | POD1 | 0.193 | 3.08 | 1.79 |
| IL-13 | POD | POD2 | 0.193 | -0.24 | 1.29 |
| IL-13 | sex | Male | 0.754 | -0.32 | 1.01 |
| IL-13 | race | Black | 0.128 | 4.00 | 2.57 |
| IL-13 | surgical duration |  | 0.717 | 0.003 | 0.01 |
| IL-13 | surgical type | pectus | 0.157 | 2.43 | 1.71 |
| IL-13 | MEQ POD0-2 |  | 0.050 | 2.44 | 1.24 |
| IL-1B | log_cytokine_AUC |  | 0.014 | 1.43 | 0.58 |
| IL-1B | POD | POD1 | 0.037 | 4.58 | 2.48 |
| IL-1B | POD | POD2 | 0.037 | -1.77 | 1.42 |
| IL-1B | sex | Male | 0.788 | 0.26 | 0.98 |
| IL-1B | race | Black | 0.298 | 2.49 | 2.39 |
| IL-1B | surgical duration |  | 0.958 | 0.0004 | 0.01 |
| IL-1B | surgical type | pectus | 0.051 | 3.53 | 1.79 |
| IL-1B | MEQ POD0-2 |  | 0.005 | 4.49 | 1.57 |
| IL-1ra | log_cytokine_AUC |  | 0.667 | 0.53 | 1.20 |
| IL-1ra | POD | POD1 | 0.471 | 1.62 | 2.21 |
| IL-1ra | sex | Male | 0.343 | -2.78 | 2.85 |
| IL-1ra | race | Black | 0.114 | 8.15 | 4.96 |
| IL-1ra | surgical duration |  | 0.448 | 0.01 | 0.01 |
| IL-1ra | surgical type |  |  | . | . |
| IL-1ra | MEQ POD0-2 |  | 0.577 | 0.90 | 1.59 |
| IL-2 | log_cytokine_AUC |  | 0.098 | 0.95 | 0.57 |
| IL-2 | POD | POD1 | 0.209 | 1.66 | 1.87 |
| IL-2 | POD | POD2 | 0.209 | -1.79 | 1.44 |
| IL-2 | sex | Male | 0.976 | 0.03 | 1.00 |
| IL-2 | race | Black | 0.191 | 3.39 | 2.55 |
| IL-2 | surgical duration |  | 0.615 | 0.004 | 0.01 |
| IL-2 | surgical type | pectus | 0.092 | 2.94 | 1.73 |
| IL-2 | MEQ POD0-2 |  | 0.052 | 2.38 | 1.21 |
| IL-4 | log_cytokine_AUC |  | 0.809 | -0.11 | 0.44 |
| IL-4 | POD | POD1 | 0.216 | 3.21 | 1.89 |
| IL-4 | POD | POD2 | 0.216 | -0.16 | 1.33 |
| IL-4 | sex | Male | 0.732 | -0.35 | 1.00 |
| IL-4 | race | Black | 0.133 | 3.96 | 2.58 |
| IL-4 | surgical duration |  | 0.731 | 0.003 | 0.01 |
| IL-4 | surgical type | pectus | 0.176 | 2.36 | 1.73 |
| IL-4 | MEQ POD0-2 |  | 0.037 | 2.59 | 1.23 |
| IL-5 | log_cytokine_AUC |  | 0.799 | 0.12 | 0.46 |
| IL-5 | POD | POD1 | 0.050 | 6.02 | 2.50 |
| IL-5 | POD | POD2 | 0.050 | -0.04 | 1.41 |
| IL-5 | sex | Male | 0.993 | -0.01 | 1.00 |
| IL-5 | race | Black | 0.169 | 3.36 | 2.43 |
| IL-5 | surgical duration |  | 0.912 | 0.001 | 0.01 |
| IL-5 | surgical type | pectus | 0.114 | 2.95 | 1.85 |
| IL-5 | MEQ POD0-2 |  | 0.008 | 4.34 | 1.61 |
| IL-6 | log_cytokine_AUC |  | 0.392 | -0.30 | 0.34 |
| IL-6 | POD | POD1 | 0.146 | 3.60 | 1.87 |
| IL-6 | POD | POD2 | 0.146 | 0.21 | 1.41 |
| IL-6 | sex | Male | 0.781 | -0.28 | 1.00 |
| IL-6 | race | Black | 0.127 | 3.97 | 2.54 |
| IL-6 | surgical duration |  | 0.760 | 0.002 | 0.01 |
| IL-6 | surgical type | pectus | 0.204 | 2.19 | 1.71 |
| IL-6 | MEQ POD0-2 |  | 0.041 | 2.52 | 1.22 |
| IL-7 | log_cytokine_AUC |  | 0.006 | 1.97 | 0.71 |
| IL-7 | POD | POD1 | 0.013 | 4.34 | 2.47 |
| IL-7 | POD | POD2 | 0.013 | -2.89 | 1.60 |
| IL-7 | sex | Male | 0.768 | 0.29 | 0.98 |
| IL-7 | race | Black | 0.279 | 2.57 | 2.36 |
| IL-7 | surgical duration |  | 0.890 | -0.001 | 0.01 |
| IL-7 | surgical type | pectus | 0.032 | 3.91 | 1.80 |
| IL-7 | MEQ POD0-2 |  | 0.006 | 4.35 | 1.55 |
| IL-8 | log_cytokine_AUC |  | 0.785 | -0.09 | 0.35 |
| IL-8 | POD | POD1 | 0.201 | 3.07 | 1.76 |
| IL-8 | POD | POD2 | 0.201 | -0.13 | 1.32 |
| IL-8 | sex | Male | 0.722 | -0.36 | 1.01 |
| IL-8 | race | Black | 0.129 | 4.01 | 2.59 |
| IL-8 | surgical duration |  | 0.742 | 0.003 | 0.01 |
| IL-8 | surgical type | pectus | 0.182 | 2.32 | 1.72 |
| IL-8 | MEQ POD0-2 |  | 0.038 | 2.58 | 1.23 |
| TNFα | log_cytokine_AUC |  | 0.545 | 0.56 | 0.92 |
| TNFα | POD | POD1 | 0.182 | 2.59 | 1.80 |
| TNFα | POD | POD2 | 0.182 | -1.30 | 1.99 |
| TNFα | sex | Male | 0.726 | -0.34 | 0.98 |
| TNFα | race | Black | 0.127 | 3.89 | 2.49 |
| TNFα | surgical duration |  | 0.706 | 0.003 | 0.01 |
| TNFα | surgical type | pectus | 0.114 | 2.87 | 1.80 |
| TNFα | MEQ POD0-2 |  | 0.029 | 2.66 | 1.21 |

MEQ: Morphine equivalents/kg; POD: Postoperative day

Green highlights indicate p-values <.05

Table S5. Mixed effects full model results for associations of log cytokine area under curve (longitudinal changes in cytokine concentrations over postoperative days 0-2 (POD 0-2)) (dependent variable) and chronic post-surgical pain (CPSP)

| **cytokine** | **Parameter/covariate** | **level** | **Type3_Pvalue** | **Estimate** | **StdErr** |
| --- | --- | --- | --- | --- | --- |
| CCL22 | CPSP |  | 0.069 | -0.49 | 0.26 |
| CCL22 | POD | POD1 | 0.170 | 0.26 | 0.18 |
| CCL22 | sex | Male | 0.956 | 0.02 | 0.41 |
| CCL22 | race | Black | 0.038 | -1.04 | 0.47 |
| CCL22 | surgical duration |  | 0.399 | 0.0017 | 0.0019 |
| CCL22 | surgical type |  | . | . | . |
| CCL22 | MEQ POD0-2 |  | 0.958 | -0.01 | 0.19 |
| EGF | CPSP |  | 0.938 | -0.01 | 0.14 |
| EGF | POD | POD1 | 0.016 | 0.43 | 0.16 |
| EGF | sex | Male | 0.675 | -0.09 | 0.2 |
| EGF | race | Black | 0.133 | -0.54 | 0.34 |
| EGF | surgical duration |  | 0.076 | 0.0019 | 0.001 |
| EGF | surgical type |  | . | . | . |
| EGF | MEQ POD0-2 |  | 0.159 | -0.21 | 0.14 |
| Fractalkine | CPSP |  | 0.724 | 0.08 | 0.23 |
| Fractalkine | POD | POD1 | 0.058 | 0.4 | 0.2 |
| Fractalkine | sex | Male | 0.691 | 0.14 | 0.35 |
| Fractalkine | race | Black | 0.200 | -0.55 | 0.42 |
| Fractalkine | surgical duration |  | 0.292 | 0.0018 | 0.0017 |
| Fractalkine | surgical type |  | . | . | . |
| Fractalkine | MEQ POD0-2 |  | 0.368 | -0.16 | 0.18 |
| GM_CSF | CPSP |  | 0.260 | 0.17 | 0.15 |
| GM_CSF | POD | POD1 | <.0001 | 0.83 | 0.33 |
| GM_CSF | POD | POD2 | <.0001 | 1.62 | 0.14 |
| GM_CSF | sex | Male | 0.736 | -0.06 | 0.18 |
| GM_CSF | race | Black | 0.175 | 0.56 | 0.41 |
| GM_CSF | surgical duration |  | 0.919 | 0.0001 | 0.0013 |
| GM_CSF | surgical type | pectus | 0.056 | -0.58 | 0.3 |
| GM_CSF | MEQ POD0-2 |  | 0.514 | 0.15 | 0.23 |
| IFNγ | CPSP |  | 0.544 | 0.08 | 0.13 |
| IFNγ | POD | POD1 | <.0001 | 0.65 | 0.2 |
| IFNγ | POD | POD2 | <.0001 | 1.65 | 0.11 |
| IFNγ | sex | Male | 0.260 | -0.18 | 0.16 |
| IFNγ | race | Black | 0.257 | 0.43 | 0.38 |
| IFNγ | surgical duration |  | 0.716 | -0.0004 | 0.0012 |
| IFNγ | surgical type | pectus | 0.031 | -0.58 | 0.26 |
| IFNγ | MEQ POD0-2 |  | 0.172 | 0.25 | 0.18 |
| IL-10 | CPSP |  | 0.721 | -0.07 | 0.2 |
| IL-10 | POD | POD1 | <.0001 | 0.5 | 0.38 |
| IL-10 | POD | POD2 | <.0001 | 1.57 | 0.22 |
| IL-10 | sex | Male | 0.093 | -0.4 | 0.23 |
| IL-10 | race | Black | 0.641 | 0.26 | 0.55 |
| IL-10 | surgical duration |  | 0.742 | 0.0006 | 0.0017 |
| IL-10 | surgical type | pectus | 0.069 | -0.73 | 0.4 |
| IL-10 | MEQ POD0-2 |  | 0.596 | -0.14 | 0.27 |
| IL-12P70 | CPSP |  | 0.943 | 0.01 | 0.14 |
| IL-12P70 | POD | POD1 | <.0001 | 0.49 | 0.34 |
| IL-12P70 | POD | POD2 | <.0001 | 1.46 | 0.15 |
| IL-12P70 | sex | Male | 0.288 | -0.18 | 0.17 |
| IL-12P70 | race | Black | 0.399 | 0.33 | 0.38 |
| IL-12P70 | surgical duration |  | 0.715 | -0.0005 | 0.0012 |
| IL-12P70 | surgical type | pectus | 0.013 | -0.74 | 0.29 |
| IL-12P70 | MEQ POD0-2 |  | 0.822 | -0.05 | 0.23 |
| IL-13 | CPSP |  | 0.044 | -0.47 | 0.23 |
| IL-13 | POD | POD1 | <.0001 | 1.21 | 0.25 |
| IL-13 | POD | POD2 | <.0001 | 1.64 | 0.15 |
| IL-13 | sex | Male | 0.012 | -0.68 | 0.26 |
| IL-13 | race | Black | 0.612 | 0.33 | 0.65 |
| IL-13 | surgical duration |  | 0.593 | -0.0011 | 0.002 |
| IL-13 | surgical type | pectus | 0.705 | -0.16 | 0.43 |
| IL-13 | MEQ POD0-2 |  | 0.245 | 0.29 | 0.25 |
| IL-1B | CPSP |  | 0.968 | -0.01 | 0.15 |
| IL-1B | POD | POD1 | <.0001 | 0.84 | 0.34 |
| IL-1B | POD | POD2 | <.0001 | 1.52 | 0.15 |
| IL-1B | sex | Male | 0.210 | -0.23 | 0.18 |
| IL-1B | race | Black | 0.165 | 0.58 | 0.41 |
| IL-1B | surgical duration |  | 0.816 | 0.0003 | 0.0013 |
| IL-1B | surgical type | pectus | 0.130 | -0.47 | 0.31 |
| IL-1B | MEQ POD0-2 |  | 0.797 | -0.06 | 0.24 |
| IL-1ra | CPSP |  | 0.529 | -0.21 | 0.32 |
| IL-1ra | POD | POD1 | 0.000 | 1.05 | 0.22 |
| IL-1ra | sex | Male | 0.545 | -0.31 | 0.51 |
| IL-1ra | race | Black | 0.157 | -0.86 | 0.59 |
| IL-1ra | surgical duration |  | 0.745 | 0.0008 | 0.0024 |
| IL-1ra | surgical type |  | . | . | . |
| IL-1ra | MEQ POD0-2 |  | 0.494 | -0.16 | 0.23 |
| IL-2 | CPSP |  | 0.234 | 0.18 | 0.15 |
| IL-2 | POD | POD1 | <.0001 | 0.92 | 0.27 |
| IL-2 | POD | POD2 | <.0001 | 1.75 | 0.16 |
| IL-2 | sex | Male | 0.081 | -0.31 | 0.18 |
| IL-2 | race | Black | 0.407 | -0.35 | 0.42 |
| IL-2 | surgical duration |  | 0.124 | -0.0021 | 0.0013 |
| IL-2 | surgical type | pectus | 0.007 | -0.83 | 0.3 |
| IL-2 | MEQ POD0-2 |  | 0.075 | 0.36 | 0.2 |
| IL-4 | CPSP |  | 0.411 | -0.15 | 0.19 |
| IL-4 | POD | POD1 | <.0001 | 0.53 | 0.24 |
| IL-4 | POD | POD2 | <.0001 | 1.65 | 0.14 |
| IL-4 | sex | Male | 0.028 | -0.49 | 0.22 |
| IL-4 | race | Black | 0.464 | 0.39 | 0.53 |
| IL-4 | surgical duration |  | 0.737 | -0.0005 | 0.0016 |
| IL-4 | surgical type | pectus | 0.043 | -0.72 | 0.35 |
| IL-4 | MEQ POD0-2 |  | 0.703 | 0.08 | 0.22 |
| IL-5 | CPSP |  | 0.553 | -0.11 | 0.19 |
| IL-5 | POD | POD1 | <.0001 | 0.52 | 0.42 |
| IL-5 | POD | POD2 | <.0001 | 1.54 | 0.18 |
| IL-5 | sex | Male | 0.154 | -0.33 | 0.23 |
| IL-5 | race | Black | 0.404 | 0.45 | 0.53 |
| IL-5 | surgical duration |  | 0.957 | 0.0001 | 0.0017 |
| IL-5 | surgical type | pectus | 0.083 | -0.68 | 0.39 |
| IL-5 | MEQ POD0-2 |  | 0.719 | -0.11 | 0.3 |
| IL-6 | CPSP |  | 0.058 | -0.51 | 0.26 |
| IL-6 | POD | POD1 | <.0001 | 1.92 | 0.34 |
| IL-6 | POD | POD2 | <.0001 | 2.36 | 0.21 |
| IL-6 | sex | Male | 0.013 | -0.78 | 0.31 |
| IL-6 | race | Black | 0.458 | 0.55 | 0.74 |
| IL-6 | surgical duration |  | 0.685 | 0.0009 | 0.0023 |
| IL-6 | surgical type | pectus | 0.258 | -0.57 | 0.5 |
| IL-6 | MEQ POD0-2 |  | 0.861 | -0.06 | 0.31 |
| IL-7 | CPSP |  | 0.798 | 0.03 | 0.12 |
| IL-7 | POD | POD1 | <.0001 | 0.71 | 0.25 |
| IL-7 | POD | POD2 | <.0001 | 1.65 | 0.11 |
| IL-7 | sex | Male | 0.312 | -0.14 | 0.14 |
| IL-7 | race | Black | 0.291 | 0.35 | 0.33 |
| IL-7 | surgical duration |  | 0.343 | 0.001 | 0.001 |
| IL-7 | surgical type | pectus | 0.029 | -0.54 | 0.24 |
| IL-7 | MEQ POD0-2 |  | 0.861 | 0.03 | 0.19 |
| IL-8 | CPSP |  | 0.013 | -0.62 | 0.25 |
| IL-8 | POD | POD1 | <.0001 | 0.61 | 0.31 |
| IL-8 | POD | POD2 | <.0001 | 1.83 | 0.18 |
| IL-8 | sex | Male | 0.002 | -0.9 | 0.29 |
| IL-8 | race | Black | 0.397 | 0.59 | 0.7 |
| IL-8 | surgical duration |  | 0.772 | 0.0006 | 0.0021 |
| IL-8 | surgical type | pectus | 0.227 | -0.57 | 0.47 |
| IL-8 | MEQ POD0-2 |  | 0.916 | -0.03 | 0.29 |
| TNFα | CPSP |  | 0.984 | 0 | 0.08 |
| TNFα | POD | POD1 | <.0001 | 0.62 | 0.17 |
| TNFα | POD | POD2 | <.0001 | 1.81 | 0.11 |
| TNFα | sex | Male | 0.550 | -0.06 | 0.09 |
| TNFα | race | Black | 0.989 | 0 | 0.21 |
| TNFα | surgical duration |  | 0.046 | 0.0014 | 0.0007 |
| TNFα | surgical type | pectus | <.0001 | -0.76 | 0.16 |
| TNFα | MEQ POD0-2 |  | 0.739 | 0.04 | 0.12 |

MEQ: Morphine equivalents/kg; POD: Postoperative day

Green highlights indicate p-values <.05

**Supplementary figure legend**

Supplementary Fig 1: Example of plots of cytokine levels before (upper plot) and after (lower plot) batch effects were addressed. IL-2 cytokine levels are plotted over time and batches are indicated by different colors.

Supplementary Fig 2: Raw cytokine data in subjects with chronic post-surgical pain and controls are presented over regrouped time points. Visit IDs indicate the time points of blood collection (Figure 1). NA refers to data for five subjects who were lost to follow up and could not be classified. CPSP: Chronic post-surgical pain.
